# Job Demands, Social Support, and Burnout among Public Senior High School Teachers, Ghana

**DOI:** 10.1101/2023.11.21.23298859

**Authors:** Richard Akutey, Edward Wilson Ansah, Daniel Apaak

## Abstract

This study aimed to investigate the extent to which job demands and social support predict burnout of public senior high school (S.H.S.) teachers in Ghana, and to determine the mediating effect of job resources in the relation between job demands and burnout among these teachers. Employing a quantitative survey, 1028 public S.H.S. teachers were selected using purposive and voluntary sampling methods. A questionnaire adopted from pre-existing standardized instruments yielded composite reliability between 0.94 and 0.98. Data was analyzed using mean, standard deviation, and multiple linear regression. Results revealed a high level of job demands (M = 3.23, SD = 0.43), social support (M = 3.02, SD = 0.54), and burnout (M = 3.33, SD = 0.92) among the teachers. Also, multiple linear regression results indicate that job demands, and social support predict burnout of the teachers. Furthermore, social support is a partial mediator of the effect of job demands on teacher’s burnout. Therefore, perceived high level of burnout is an effect of high levels of job demands, which poses a serious threat to the health and well-being of these teachers and compromises teaching quality in Ghana’s S.H.S. However, this challenge can be prevented or reduced by providing more social support to the teachers. Hence, government, management, and other educational stakeholders need to provide a strong safety leadership in all matters that concerns teacher’s health and safety. The school administrators and teachers are also encouraged to promote social support vertically and horizontally.

## Introduction

Every profession has its own occupational health problems, due to the kind of work performed [1]. Teaching is a stressful occupation [2], with a lot of occupational health challenges, including burnout, physical and psychosocial hazards because of high job demands [2]. Prior researches have shown that among all occupational groups, teachers are more vulnerable to psychological distress, work stress, and burnout [3, 4]. This is because teachers experience wide range of stressors, including indiscipline problems of students, teaching preparation, conflicts with colleagues, value conflicts, and role ambiguity [3]. High job demands associated with the teaching profession make teachers experience on-the-job burnout [1, 5]. Burnout encounter by teachers is not only associated with high levels of job demands, but also job resources like social support [5, 6, 7]. Social support is a form of job resources and one way of viewing it is the support obtained from coworkers and administration or employers [6, 7]. The evidence is that when there is positive social relation between fellow teachers and management of the school, teacher burnout level is lowered [2, 7].

Maintaining a safe and socially conducive work environment in schools is important for learners, teachers, and administrators [1]. The office of every teacher is the classroom, thus, efforts aimed at providing the “best” working environment for teachers cannot be of less concern. Although schoolteachers are one of the numerous professional groups in Ghana, they have not habitually been the focal of many researchers in the country.

Meanwhile, many occupational health issues such as burnout come about because of classroom teaching [5]. Such situation became worse when the government of Ghana introduced a flagship programme; the Free Senior High School (FSHS) since 2017 [8].

FSHS programme has led to an incremental jump in the enrollment of students in the senior high school (S.H.S.) level [8], of 72 percent in 2017-2018 compared to 2016-2017 academic year [8]. Consequentially, the job demands in these public S.H.S. for teachers and other school workers has increased tremendously. The result includes over burdening of Ghana educational system thereby compelling the Ghana Education Service (GES) to run a shift system called, the Double Track System (Gold and Green shift system) [9]. Evidence by Asumadu [10] on “challenges and prospects of the Ghana FSHS policy” revealed that although the policy has come to ensure increase in enrolment in public S.H.S., but the implementation is hindered by several factors with inadequate teachers being one of the major factors. Thus, such an educational policy put a tremendous burden on the Ghanaian teacher at the S.H.S. level.

Inadequacy of on teachers to match students’ enrollment had intensified the already existing occupational health problems of teachers such as burnout. Job burnout is a serious syndrome that affects the health of workers at the work environment, and World Health Organization (WHO) has gone ahead to include it in its handbook of diseases, calling for more research on it [11]. Hence, the individual and social impacts of job burnout among public S.H.S. teachers in Ghana highlights the need for immediate preventive evidence-based interventions [12]. Apparently, the well-being of teachers could be achieved when preventive interventions relating to satisfactory work are driven by evidence. Such research is crucial for teacher health and well-being that could translate into quality teaching and production of relevant 21^st^ century graduates for Ghana. Unfortunately, there seems to be no available research evidence from Ghana on the roles of high job demands and job resources on teacher burnout since the introduction of FSHS programme. Therefore, the purpose of this study was to investigate the extent to which job demands and social support predict burnout of public S.H.S. teachers in Ghana, and to determine the mediating effect of job resources in the interaction between job demands and burnout of the teachers.

## Methods

## Participants’ Selection

Employing a quantitative survey, 1028 public S.H.S. teachers were selected using purposive and voluntary sampling methods. These public S.H.S. teachers were selected because the impact of FSHS policy is felt on only public S.H.S. The participants comprised 52% (n = 537) males and 48% (n = 491) females, with age range of 24 – 55 years (M = 35, SD = 7). Sixty-five percent (n = 671) of the participants had first (bachelor’s) degree, and 35% (n = 357) master’s degree (MPhil, MSc, or MED). Moreover, 65% (n = 665) attained a rank of Principal Superintendent, 26% (n = 268) Assistant Director II, 7% (n = 75) Assistant Director I, and 2% (n = 20) Deputy Directors. The median, mean and standard deviation for years of teaching experience are seven years, nine years, and seven years respectively, with the minimum and maximum being one year and 32 years, respectively. Unfortunately, 7% (n = 74), 6% (n = 59), and 6% (n = 57) of the participants have been diagnosed of hypertension, diabetes, and other heart or heart-related conditions, respectively.

### Measures

A questionnaire, measuring burnout, job demands and social support, was used to collect data from the participants. Burnout items were adopted from the Maslach Burnout Inventory – Educators Survey (MBI-ES) [13], which measured; a) emotional exhaustion, b) depersonalization, and c) personal achievement. Some of the items were: *“I feel emotionally drained from my work*.*”, “I’ve become more heartless toward people since I took this job*.*”*, and “*I don’t really care what happens to some students*.*”* respectively. Responses were on a 7-point Likert scale from 0 (Never) to 6 (Everyday), with higher scores signaling high level of burnout of the teachers.

Social support items were derived from Social Provisions Scale [4, 5], which measured two sub-variables: (a) collegial support, and (b) supervisory support. Some of the items were: *“In educational matters, I can always get good help from my colleagues”*, and “*In educational matters, I can always get help and advice from the school leadership or management”* respectively. Job demands items were also adopted from Job Content Questionnaire [5, 14] which measured two sub-variables: (a) time pressure, and (b) discipline challenges at school. Some of the items included: “*Preparation for teaching must often be done after working hours”* and *“My teaching is often disrupted by students who lack discipline”* respectively. Responses for both social support and job demands were on a 4-point Likert scale, ranging from “Strongly Disagree” (1) to “Strongly Agree” (4). A high overall score indicate high level of social support and job demands.

These pre-existing instruments have been used by many researchers and they recorded high and acceptable reliability and validity values across nations and worker segments [5, 13, 15, 16, 17]. The questionnaire yielded overall Cronbach alpha reliability of 0.90, with 0.77, 0.87, and 0.91 for job demands, social support, and burnout constructs, respectively. The composite reliability indices of job demand (0.94), social support (0.98), and burnout (0.97) were also acceptable. Additionally, the questionnaire yielded high and acceptable convergent validity values; with job demand = 0.68, social support = 0.72, and burnout = 0.60. The questionnaire also measured participants’ socio-demographic characteristics such as gender, age, highest educational level, rank, years of teaching experience, status of hypertension, diabetes, and other heart or heart-related conditions.

### Procedures

The University of Cape Coast Institutional Review Board (UCCIRB) issued ethical clearance (UCCIRB/CES/2022/141), and GES also granted the required approvals for this study. Data collection was on online with electronic questionnaire, designed into Google Form where the link (in URL) was shared on WhatsApp platforms of the teachers. One-month duration was given to the teachers to fill and submit their responses. To prevent multiple responses, the introductory page of the Google Form indicated that “the questionnaire MUST be filled ONLY ONCE and NEVER again” though it will be shared on the platforms multiple times. Moreover, it was clearly stated the “Questionnaire is for ONLY teachers in S.H.S. in the Eastern Region of Ghana”. The link was blocked after expiration of data collection. The data collection took place from 1^st^ October, 2022 through to 31^st^ December, 2022. Participants were assured of their anonymity, confidentiality of their information and voluntary participation in the study, and that they could stop answering the questionnaire at any time without a penalty. Teachers who agreed to participate in the study were required to properly check a box on the questionnaire’s introduction page to confirm their willingness to participate in this study.

## Data Analysis

The level of social support, job demand, and burnout were each aggregated as a single score from eight (8), eight (8) and twenty-two (22) items, respectively. Using benchmarks of 1 to 4 for social support and job demand, and 0 to 6 for burnout, participants’ rating in terms of the mean scores was reported as ≤ 1.9 = low, 2.0 – 2.9 = moderate, and ≥ 3.0 = high for job demands and social support. While ≤ 1.5 = low, 1.6 – 3.0 = moderate, and ≥ 3.1 = high for burnout. The data satisfied all the necessary validity and reliability criteria such as composite reliability, convergent and discriminant validity [18]. Besides, the constructs recorded composite reliability of 0.70 or higher. Furthermore, the indicators and constructs’ AVE of 0.60 or higher were acceptable [18, 19, 20].

## Result

The results indicate high level of job demands on the teachers (M = 3.23, SD = 0.43), as a results of time pressure (M = 3.29, SD = 0.47) and students’ discipline challenges (M = 3.13, SD = 0.62), respectively. Also, social support was high (M = 3.02, SD = 0.54), coming from collegial support (M = 3.02, SD = 0.59) and supervisory support (M = 3.02, SD = 0.69). Furthermore, burnout level was high (M = 3.33, SD = 0.92) due to high levels of emotional exhaustion (M = 3.53, SD = 1.18), and personal accomplishment (M = 3.72, SD = 1.07), and medium level of depersonalization (M = 2.86, SD = 0.95).

Further analyses using multiple linear regression analysis indicated item loadings are between 0.703 and 0.933 on job demands, 0.776 and 0.907 on social support, and 0.554 and 0.854 on burnout. These loadings are high, and that the items loaded highest on their respective latent variables than they did on other variables (see Table 1), indicating item reliability [21, 22]. Also, inner model collinearity showed no such issues between the constructs, thus, the variance inflation factor (VIF) for both job demand and social support (IVs) was 1.089 (see Table 1). Additionally, composite reliability and convergent validity were fulfilled [20, 23, 24]. Moreover, using Fornell and Larcker [25] criterion, job demands, social support and burnout achieved acceptable discriminant validity (see Table 2).

**Table 1.**
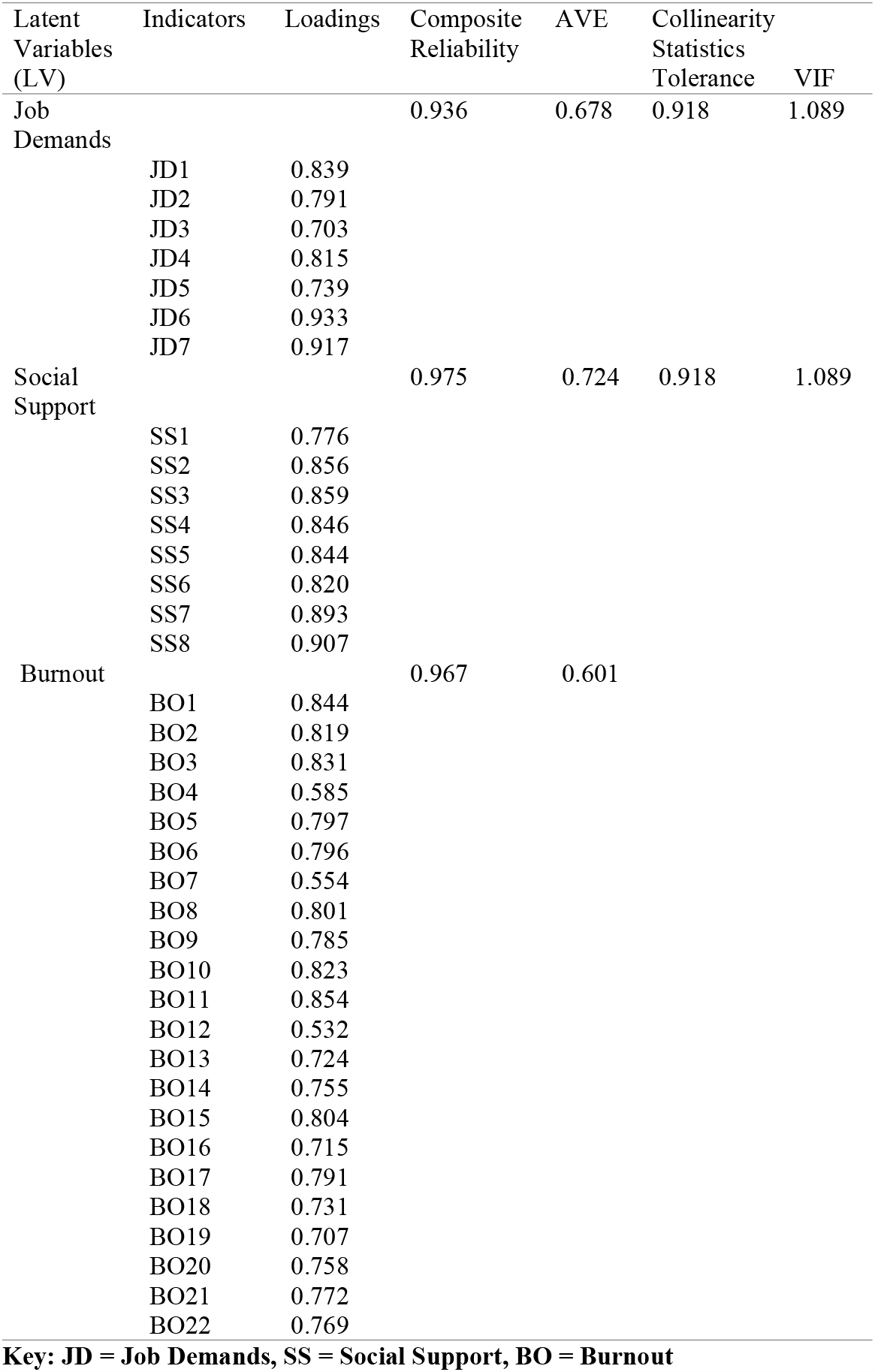
Summary of the Results of Reflective Model l and Multicollinearity Diagnostic.

**Table 2:** Discriminant Validity using Fornell and Larcker Criterion.

| Latent Variables | BO | JD | SS |
| --- | --- | --- | --- |
| BO | **0.775** |  |  |
| JD | 0.555 | **0.823** |  |
| SS | 0.457 | 0.511 | **0.851** |

**Key: JD = Job Demands, SS = Social Support, BO = Burnout**

The analysis further indicates that the combination of job demands, and social support explained 12% of the variance in burnout (R^2^ adj = 0.119) and hence, predicted burnout, F (2, 1025) = 70.341, P < 0.001. It is also found that there is a meaningful, positive, and low-level relation between job demands and social support, and burnout, r = 0.347; p < 0.001. Accordingly, the more job demands increases, the more burnout increases, whereas the more social support increases, the more burnout decreases. Moreover, job demands, t (2, 1025) = 10.253, p < 0.001, influence more the change in burnout among the teachers compare with social support, t (2, 1025) = 2.774, p < 0.01.

The mediating role of social support indicated a positive association with job demands, yielding combined information to later test the indirect effect (a path: B = 0.359), t (1, 1026) = 9.583, p < 0.001. Moreover, job demands was positively related to burnout (c path: B = 0.719), t (1, 1026) = 11.494, p < 0.001. Lastly, social support was positively associated with burnout (b path: B = 0.297), t (1, 1026) = 5.682, p < 0.001. Bootstrapping further confirmed the mediating role of social support in the relationship between job demands and burnout of the teachers (B = 0.144; CI = 0.0330 to 0.257). In addition, results indicated that the direct effect of job demands on burnout remained significant (c’ path: B = 0.144), t (1, 1026) = 2.774, p < 0.01 when controlling for social support, thus, suggesting a partial mediation role of social support in reducing the effect of job demands on burnout among the teachers [6, 26] (see Figure 1) (see Table 3).

**Table 3:**
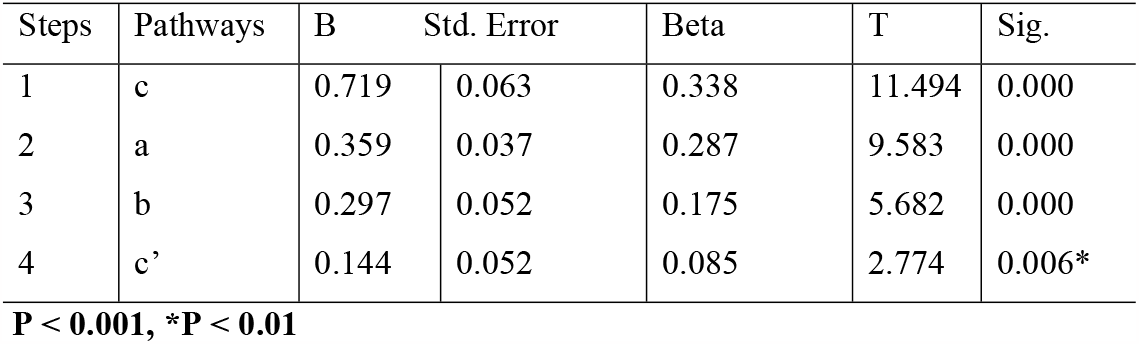
Mediation Steps for Job Demands and Social Support.

**Figure 1.**
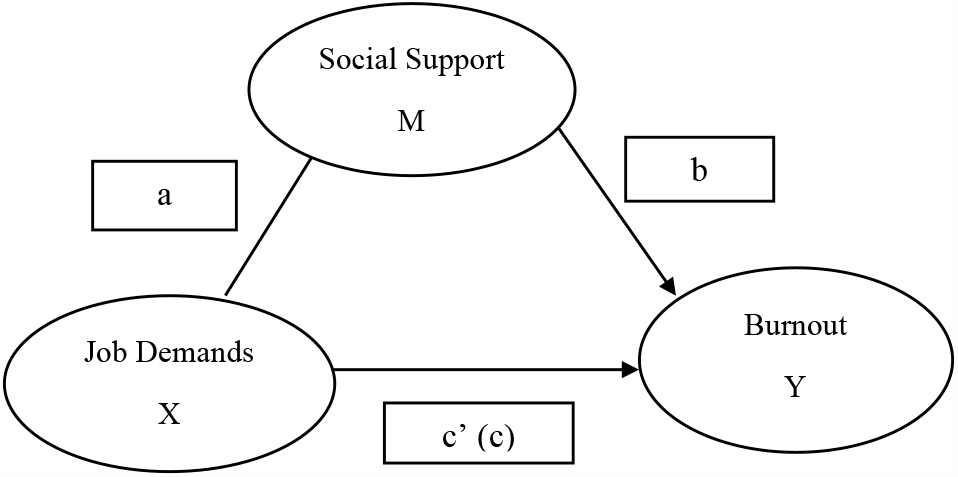
Mediation Pathway of Social Support on the Effect of Job Demands on Burnout.

## Discussion

The findings reveal that S.H.S. teachers in the Eastern Region of Ghana perceived their job to be highly demanding, as a result of high time pressures (including workloads) and students’ discipline challenges in their various schools. This is consistent with the previous research findings [10, 14], which have also attributed high job demand of teachers to high time pressure (workload) where teachers have little to no time for relaxation and rehabilitation. However, this situation may be aggravated in the face of exponential increase in students’ enrollment due to the implementation of FSHS policy without the corresponding employment of new teachers [10]. These under-pressured teachers combine classroom teaching and paperwork with other workloads such as involvement in numerous school development projects, frequent meetings, constant changes, and engagement with parents [27]. This high job demands associated with the teaching profession make teachers suffer various forms of injuries and ill health from burnout.

The finding further revealed that teachers experience burnout and exhibit high level of exhaustion. This is consistent with the finding from Sokal, Trudel, and Babb [28], that high job demands is causing teachers to exhibit high level of exhaustion which may slow down their work input and output, compromising quality teaching. Besides, some key antecedents to burnout among teachers include systemic modifications in educational systems, poor working environment, administrative issues, time pressure, and students’ misconducts [27, 29]. Thus, the call for paying a particular attention to the increasing demands on teachers and associated consequences is warranted especially when a new policy is crudely introduced without attendant employment of teachers. Thus, teachers’ levels of burnout is steadily increasing [30], and this is highly contagious and that if it is not addressed pragmatically, it might spread to other teachers like a virus [30], and lower teacher health and well-being, that affect quality teaching.

The findings again showed that these teachers perceived high levels of social support from their colleagues and supervisors, may be because of the helps they do receive from their colleagues and supervisors. This is consistent with other research findings [31, 32], that teachers receive support from and share problems or good news with their work colleagues and supervisors. However, such work atmosphere may be eroded if active and pragmatic steps are not taken to increase and maintain both vertical and horizontal social supports. Additionally, the findings revealed that social support significantly predicts burnout, by reducing the negative effect of burnout on teachers. This finding is in line with and lends credence to other studies that supportive social relationships with coworkers and superiors are linked to lower levels of burnout [5, 6]. Perhaps, it would be appropriate to encourage heads and other administrators to build and maintain some high levels of supporting work atmosphere.

The finding further reveals that social support plays a partial mediation role to lower the effect of job demands on burnout among the teachers. That is to say that, though there is a high level of social support in the schools, it could cancel out the effect of high level of burnout felt by the teachers in FSHS regime. This is in line with previous research evidence [33] that social support buffers the impact of work pressures on burnout. This suggests that when teachers receive greater social support, their ability to cope with the demanding nature of teaching and related activities improves, lowering their risk of burnout. Fortunately, teachers in this study seem to have some level of both horizontal and vertical social support which could serve as a buffer to their intense level of burnout. Nevertheless, the effects of such level of social support may not be effective to counteract teacher burnout because of other serious antecedent factors. Thus, with the introduction of the FSHS and attendant increase in students’ enrollment, the workload of teachers exponentially increased complicating teacher burnout. This is most likely to result in complicated health and well-being of the teachers, that compromises teaching quality resulting in lower level of education standards. Hence, in such a school environment like public S.H.S., administrators and teachers need to engage more in sharing their work, ideas, and give listening ears and encouragement to each other, a situation that is likely to ameliorate the burden of work overload and lower burnout syndrome. Such educational setting would promote quality teaching and learning that lead to the production of 21^st^ century graduates that can meaningfully make a positive contribution to the nation’s socioeconomic progress.

Although this is the first study on job demands, social support, and burnout among public S.H.S. teachers in Ghana, post FSHS, it still has certain drawbacks. Since only educators from the Eastern Region participated, the findings of this study may understate the degree of workload, social support, and burnout at the public S.H.S level. Additionally, it may be difficult to extrapolate the results and conclusions because the participants were selected using purposive and voluntary sampling methods. However, the robust statistics used ensure the validity and dependability of the findings and conclusions.

## Conclusions

Findings from this study reveals that S.H.S. teachers in the Eastern Region of Ghana perceived their job to be highly demanding, as result of high time pressures (including workloads) and students’ discipline challenges such teachers face in their various schools. It is further revealed that teacher burnout is high because they exhibit high level of exhaustion which may compromise their health, well-being, and teaching quality. Again, these teachers perceived high levels of social support from their colleagues and supervisors, may be because of the helps they do receive from their colleagues and supervisors. Additionally, it has been revealed that social support significantly predicts burnout, by reducing the negative effect of burnout on the teachers. This means that, the more job demands increases, the more burnout increases, whereas the more social support increases, the more burnout decreases. It has also been revealed that social support plays a partial mediation role to lower the effect of job demands on burnout among the teachers. This suggests that when teachers receive greater social support, their ability to cope with the demanding nature of teaching and related activities improves, lowering their risk of burnout.

Hence, policy makers in Ghana are encouraged to prioritize addressing the issue of teacher burnout by focusing on quality improvement in the teaching profession. Thus, it is essential that GES, administrators (headmasters and headmistresses) of public S.H.S., teachers, and teachers’ unions like Concern Collation of Teachers (CCT), National Association of Graduate Teachers (NAGRAT), and Ghana National Association of Teachers (GNAT) ensure the initiation of burnout management programmes, increase in emotional and psychological support, encourage educators to support one another at work. Additionally, it is important that educational leaders in Ghana be conscious of the welfare of the teachers, support them at work, and act as the unifier by effectively getting all the teachers to coexist amicably. Moreover, introduction of one stream teaching to enable teachers to have time for resting and recovery, engagement of additional teachers to lessen the burden of those on the field, and proper stress management training before and on-the-job are important. It is also necessary to investigate how job demands, social support, and burnout vary over time since changes in these workplace critical elements have impact on teachers’ well-being and quality teaching.

## Data Availability

The datasets generated and analyzed during the current study are available from the corresponding author on reasonable request.

## Declarations

### Ethical Approval and Consent to participate

The University of Cape Coast Institutional Review Board (UCCIRB) issued ethical clearance (UCCIRB/CES/2022/141), and GES also granted the required approvals for this study. Informed consent was obtained from all participants prior to data collection.

### Consent for publication

Not applicable.

### Authors’ contributions

EWA and RA conceptualised and designed the study. RA collected the data from the participants. EWA, RA, and DA analyzed the data. RA wrote the initial draft. EWA and DA critically revised the manuscript for its intellectual content. All authors critically revised the manuscript and approved of its final version for submission.

## Acknowledgements

Not applicable

